# A structured course of disease dataset with contact tracing information in Taiwan for COVID-19 modelling

**DOI:** 10.1101/2024.02.28.24303518

**Authors:** Yu-Heng Wu, Torbjörn E. M. Nordling

## Abstract

**Background:** The COVID-19 pandemic has flooded open databases with population-level data. However, individual-level structured data, such as the course of disease and contact tracing information, is almost non-existent in open databases.

**Aim:** Publish a structured and cleaned COVID-19 dataset with the course of disease and contact tracing information for easy benchmarking of COVID-19 models.

**Methods:** We gathered data from Taiwanese open databases and daily news reports. The outcome is a structured quantitative dataset encompassing the course of the disease of Taiwanese individuals, alongside their contact tracing information.

**Results:** Our dataset comprises 579 confirmed cases covering the period from January 21, to November 9, 2020, when the original SARS-CoV-2 virus was most prevalent in Taiwan. The data include features such as travel history, age, gender, symptoms, contact types between cases, date of symptoms onset, confirmed, critically ill, recovered, and dead. We also include the daily summary data at population-level from January 21, 2020, to May 23, 2022.

**Conclusions:** Our data can help enhance epidemiological modelling.

## 1 Background & Summary

Current epidemiological models rely on population-level data, demanding precise measurements of infected individuals and deaths. However, the accuracy of reported infection numbers can be compromised due to limited testing. In Stockholm, Sweden, Roxhed *et al*. found a seroprevalence of 12.5%, implying a significant discrepancy between the potential 150,000 infections and the 10,000 PCR-confirmed cases reported during the study period. Estimating mortality rates is also challenging. The WHO reported approximately 15 million excess deaths by December 2021, compared to 5.4 million confirmed COVID-19 deaths. The Economist’s estimates varied from 15 million to 25.2 million excess deaths, far exceeding reported COVID-19 deaths. Additionally, calculating the basic reproduction number (*R*_0_) is challenging due to reporting errors in infection numbers. For COVID-19, *R*_0_ estimates ranged widely from 0.17 to 4.5 with broad confidence intervals^1^. These issues highlight the unreliability of commonly used population-level measurements.

To mitigate the inaccuracies inherent in population-level data, utilizing more informative individual-level data is advisable. For example, Lee *et al*. published a case report of the course of disease of a 46-year-old woman diagnosed based on positive real-time RT-PCR test from oropharyngeal swab samples^2^. The days of fever, cough, dyspnea, dysuria, *etc* are listed. With three consecutive negative results of real-time RT-PCR tests for SARS-CoV-2, the woman was discharged after 24 days of hospitalization. In another instance, the report on France’s first three cases^3^ meticulously documented essential details, including travel history, symptom onset, medical consultations, hospitalization, confirmation, critical condition, and contact tracing.

Individual-level data finds valuable application in predicting specific risks. Garibaldi *et al*. developed the COVID-19 Inpatient Risk Calculator (CIRC), forecasting severe illness or death within 7 days for COVID-19 patients based on data from 832 consecutive admissions^4^. Wongvibulsin *et al*. created a system predicting progression from moderate to severe illness or death within 14 days using demographics, admission source, comorbidities, vital signs, lab measurements, and clinical severity data from 3494 COVID-19 patients^5^. In the pandemic’s early stages, when individual-level data accessibility was limited, Lichtner *et al*. trained a model on 718 non-COVID viral pneumonia patients’ data, effectively predicting adverse outcomes in critically ill COVID-19 patients^6^. Unfortunately, none of these datasets are publicly available.

In this study, data is collected from diverse open-source databases, resulting in a comprehensive structured dataset encompassing demographics, epidemiological details, disease progression information, and subject contact tracing. Previously, we developed a model for forecasting the end of a local outbreak based on the moving average ratio of daily infected to suspected cases which utilizes the daily summary data we included in this dataset^7^.

## 2 Methods

We collected individual-level COVID-19 data from public Taiwanese databases and converted the data into a unified format. The data collection process and data preprocessing are described in the following sections.

### 2.1 Data collection

The data was collected from the following open-source databases: Taiwan Centers for Disease Control press release (CDC press release)^8^, United Daily News (COVID-19 Visualization)^9^, Taiwan CDC Open Data Portal, Regents of the National Center for High-performance Computing (COVID-19 Dashboard)^10^, Taiwan Centers for Disease Control open data portal (CDC open data portal)^11^, and Taiwan Centers for Disease Control press conference (CDC press conference)^12^. The flow chart of the data collection process is shown in Figure 1.

**Figure 1.**
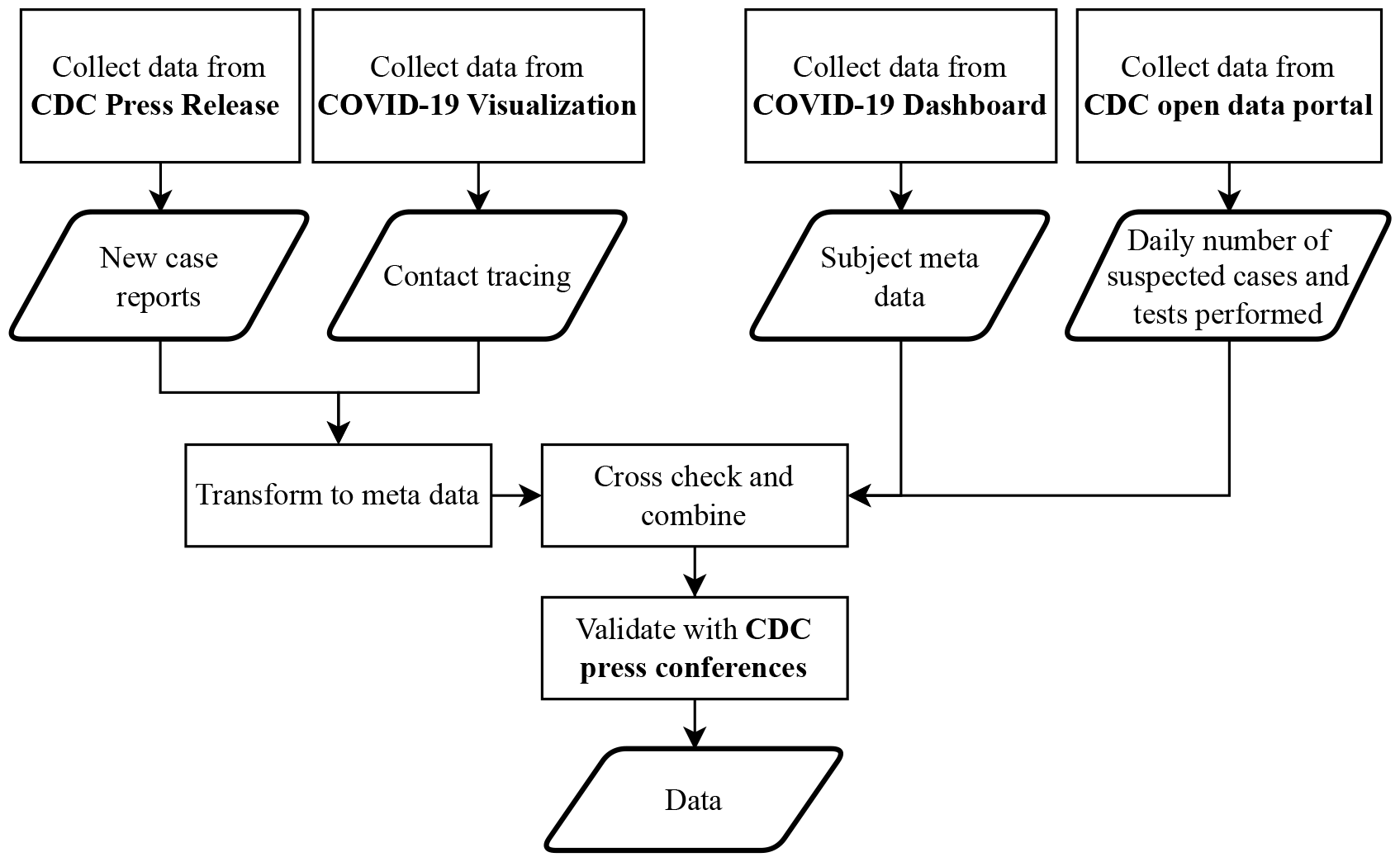
Flowchart depicting the collection process for Taiwanese COVID-19 data, including daily summary data, the course of the disease for confirmed cases, and contact information.

#### 2.1.1 Individual-level data

We collected individual-level epidemiological data, course of disease data, and contact tracing data from January 21st, 2020 to November 9th, 2020 covering the first outbreak in Taiwan. The individual-level data in our dataset primarily originates from daily reports issued by the Taiwan CDC. These reports contained detailed information about confirmed COVID-19 cases. While this source was informative, the data was presented in an unstructured text format, necessitating manual conversion into a structured and quantifiable format. We also obtained valuable data from the COVID-19 Visualization, which provided visualizations of the contact networks among locally confirmed cases. Furthermore, the COVID-19 Dashboard served as a source for the course of disease data, including report dates and confirmed dates. We augmented this information with metadata and conducted validation processes based on CDC press conference.

The confirmed cases data contained 579 samples with 64 features including travel history, age, gender, nationality, the onset of symptom, confirmed date, symptoms, way of discovery, and contact types between cases. We categorized the contact type into groups such as couple, parents, grandparents, siblings, family, friends, live together, flight, flight (nearby seat), travelling, school, car, coworker, hospital, hotel, Panshi combat ship (military ship), Coral Princess (cruise ship), and others. Some cases also included ICU admission, recovery, and death dates, as available from our sources. When accessible, Taiwan CDC provided details like the count of close contacts, contact dates, and case indexes for contacts.

#### 2.1.2 Daily summary data

The daily summary data, sourced from Taiwan CDC press releases^8^, provides essential population-level information. However, inconsistencies in reporting intervals and changes in counting methods, notably on 2020-03-06, impact data accuracy. To mitigate this, we accessed the “Daily Number of Cases Suspected SARS-CoV-2 Infection Tested” dataset from the Taiwan CDC Open Data portal^11^. This dataset comprises daily counts of suspected SARS-CoV-2 cases derived from three sources: notifications of infectious diseases, home quarantine and inspection, and expanded monitoring.

The data spans from January 21, 2020, to May 23, 2022, providing a comprehensive overview of the pandemic’s progression. To ensure data consistency and reliability, we manually transformed this data into a structured format. This dataset includes daily statistics on various COVID-19 aspects, such as the number of suspected cases, excluded cases, abroad positive cases, local positive cases, positive cases with unknown sources, deaths, recovery cases, and hospital quarantine cases.

### 2.2 Data preprocessing

When structuring the data, we identified a few anomalous cases, which we investigated as part of our data preprocessing. For instance, Case ID 19 was confirmed on February 15th, 2020. However, due to a mistaken diagnosis of pneumonia, ID 19 was placed in the ICU on February 3rd, 2020, which was before the confirmed date. Since there was no hospital transmission, it’s likely that the case was isolated after being admitted to the ICU. Therefore, the confirmed date (monitor isolation) should be readjusted to February 3rd, 2020. Case ID 530 was later identified as a false positive and subsequently removed from the dataset by the Taiwan CDC. Case ID 555 had contact with infected individuals on September 13, 2020, and was confirmed as a COVID-19 case on September 18, 2020, in the Philippines. Subsequently, the case was discharged from the hospital on October 3, 2020, tested negative on October 9, 2020, and travelled to Taiwan on October 15, 2020. The case was tested and confirmed on October 30, 2020, which resulted in a long period of susceptibility to infection. To rectify this, we removed case ID 555’s infection date.

## 3 Data Records

The data is publicly available in figShare repository^13^.

### 3.1 Data description of individual-level data

Our dataset encompasses a total of 578 cases, including epidemiological data, course of disease data, and contact tracing data. In this section, we offer a comprehensive overview of the dataset’s characteristics and composition. Table 1 provides an overview of epidemiological data, categorizing it based on case origin, travel date, travel history, gender, and age. Each category lists the number of cases alongside their corresponding percentages. Additionally, Table 2 explores the distribution of the course of disease data, indicating data availability through the count of cases with available data, along with the respective percentages. Specific aspects, such as asymptomatic date, symptoms, symptomatic date, confirmed date, critically ill date, recovery date, death date, and report to CDC date, are highlighted. Among 578 subjects, 452 had reported symptoms recorded by Taiwan CDC, and 442 of them provided their symptomatic dates. The number of recovered and deceased cases is reported in CDC’s daily summary data. The number of critically ill cases is not directly provided by Taiwan CDC and is thus estimated as the sum of critically ill and deceased cases.

**Table 1.**
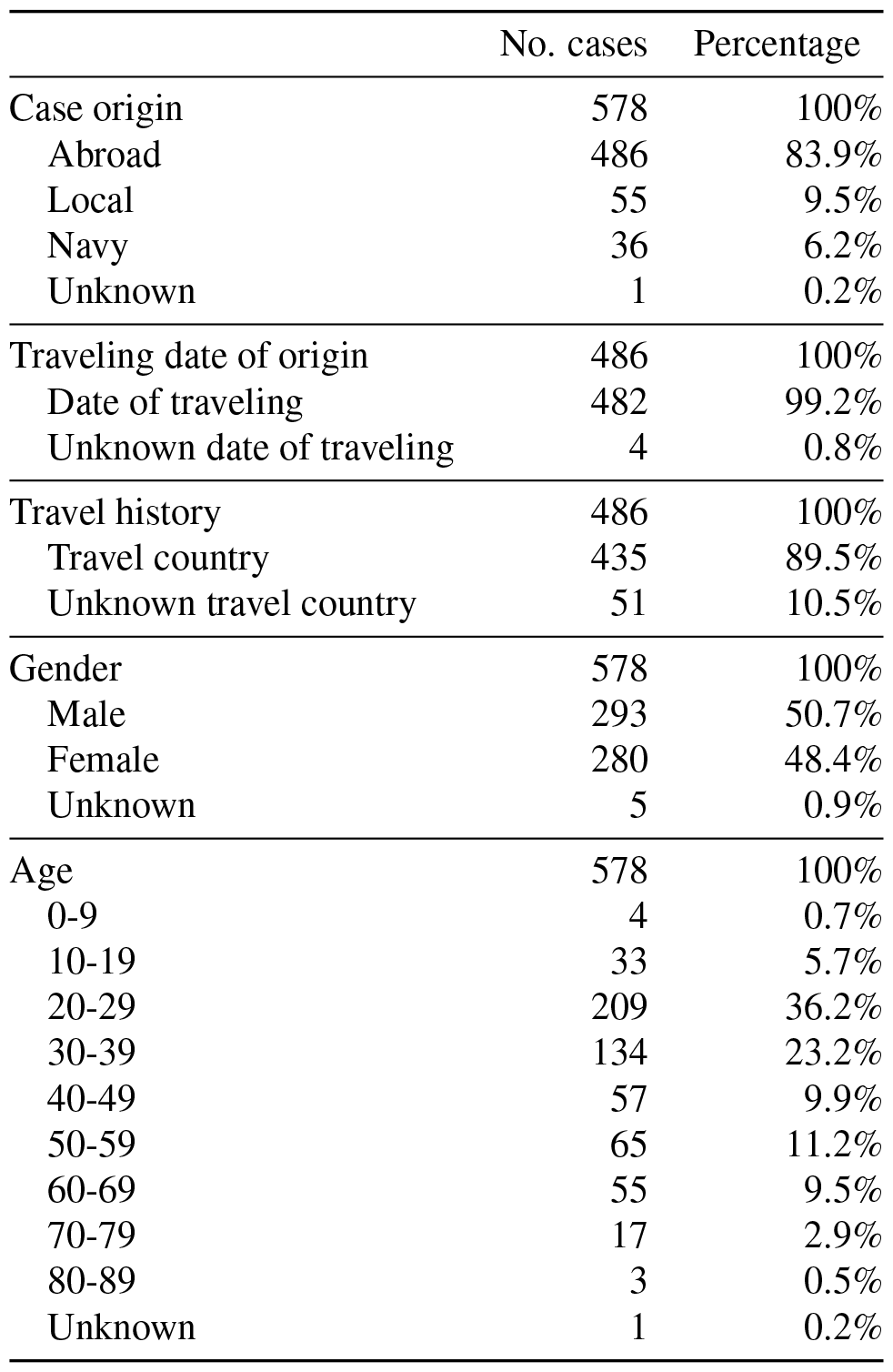
Description of epidemiological data. This table outlines the distribution of epidemiological data within the Taiwanese dataset, categorizing it by case origin, travel history, gender, and age. The dataset comprises a total of 578 cases. For each category, both the number of cases and their respective percentages are provided.

|  | No. cases | Percentage |
| --- | --- | --- |
| Case origin | 578 | 100% |
| Abroad | 486 | 83.9% |
| Local | 55 | 9.5% |
| Navy | 36 | 6.2% |
| Unknown | 1 | 0.2% |
| Traveling date of origin | 486 | 100% |
| Date of traveling | 482 | 99.2% |
| Unknown date of traveling | 4 | 0.8% |
| Travel history | 486 | 100% |
| Travel country | 435 | 89.5% |
| Unknown travel country | 51 | 10.5% |
| Gender | 578 | 100% |
| Male | 293 | 50.7% |
| Female | 280 | 48.4% |
| Unknown | 5 | 0.9% |
| Age | 578 | 100% |
| 0-9 | 4 | 0.7% |
| 10-19 | 33 | 5.7% |
| 20-29 | 209 | 36.2% |
| 30-39 | 134 | 23.2% |
| 40-49 | 57 | 9.9% |
| 50-59 | 65 | 11.2% |
| 60-69 | 55 | 9.5% |
| 70-79 | 17 | 2.9% |
| 80-89 | 3 | 0.5% |
| Unknown | 1 | 0.2% |

**Table 2.** Description of course of disease data. The table outlines the distribution of course of disease data within the Taiwanese dataset. Data availability is indicated by the number of cases with available data and the corresponding percentage. The asymptomatic date refers to the first contact date with the source case, i.e., the infection date.

|  | No. cases with available data/No. expected cases | Percentage |
| --- | --- | --- |
| Course of disease data |  |  |
| Asymptomatic date | 33/578 | 5.7% |
| Symptoms | 452/452 | 100% |
| Symptomatic date | 442/452 | 97.8% |
| Confirmed date | 578/578 | 100% |
| Critically ill date | 58/65 | 89% |
| Recovery date | 79/526 | 15% |
| Death date | 7/7 | 100% |
| Report to CDC date | 500/578 | 86.5% |

### 3.2 Kaplan-Meier plot

The Kaplan-Meier plot is a statistical tool used to estimate and visually represent the probability of an event occurring over time, commonly applied in medical studies to illustrate survival or state transition dynamics. In a subject’s course of disease, a subject could go to asymptomatic (infection with no symptoms, I^A^), symptomatic (infection with symptoms, I^S^), confirmed (C) critically ill (ICU admission, I^C^), recovered (two consecutive negative PCR tests, R), and death (D) as shown in the bottom right plot in Figure 2. Infection and immune status can be determined by an antigen test, PCR test, and antibody test. The nonpharmaceutical interventions (NPI) inhibit the transition from S to I, and the intensive care units (Care) activate the transition from I^C^ to R, and the vaccination activates the transition from S to R. In Figure 2, the Kaplan-Meier plot illustrates state transitions from one state to another without passing through intermediate states. For example, the samples from symptomatic to recovered do not include individuals who transitioned from symptomatic to critically ill to recovered.

**Figure 2.**
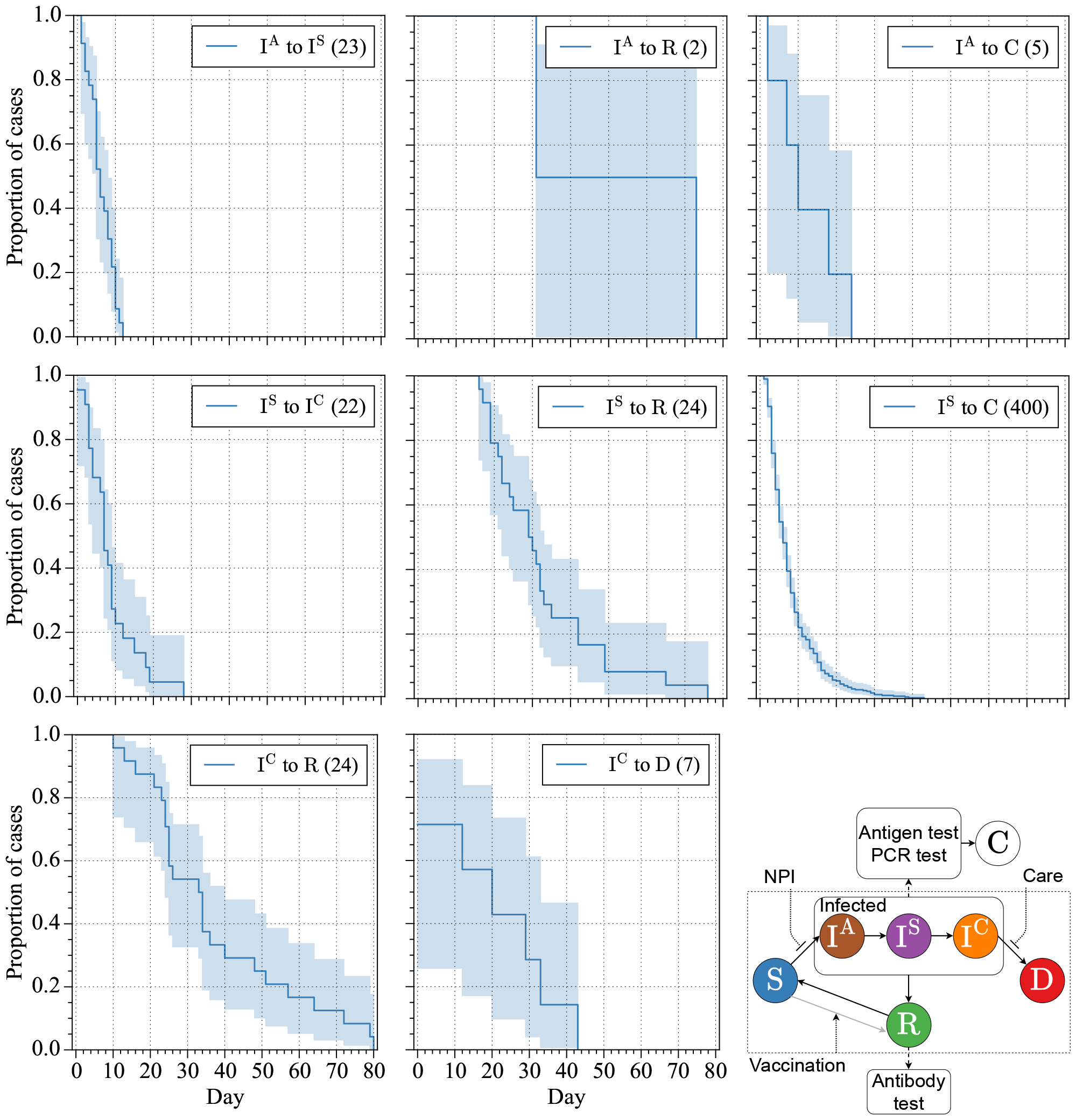
Daily state transition Kaplan-Meier plot of Taiwanese data. The 95% confidence interval was calculated by Greenwood’s Exponential method. The different states are denoted as asymptomatic (I^A^), symptomatic (I^S^), confirmed (C) critically ill (I^C^), recovered (R), and death (D). The bottom right figure depicts the state trasition process. The detail of the state transition process is described in the Supplementary.

### 3.3 Contact network

The contact network comprises 8,842 nodes, with 578 nodes representing infected individuals, constituting approximately 7% of the total nodes. In terms of edges, the network encompasses a total of 46,008 connections, of which 1,183 (2.6% of the edges) are infection-related. Among these edges, 37 have identifiable infection paths, resulting in directed edges, while the remaining 1,146 are forming undirected edges. The proportion of directed edges to total edges is 0.08%, rising to 3.1% for infection-related edges. Additionally, the network displays 152 clusters, ranging in size from 2 to 1,898 nodes, with a median cluster size of 16.5 and a mean cluster size of 58.2.

The left panel of Figure 3 presents a mixed graph representation of a contact network, where infection paths are denoted by arrows. The contact network is visualized with Cytoscape. In this instance, the initial case, I277, transmits the infection to individuals I269 and I288, and subsequently, I269 spreads the infection to I299. Distinct contact types such as ‘family’ (highlighted in red), ‘same flight’ (depicted in blue), and ‘other/unknown’ connections (in gray) are visually differentiated. Smaller nodes represent uninfected contacts within the network.

**Figure 3.**
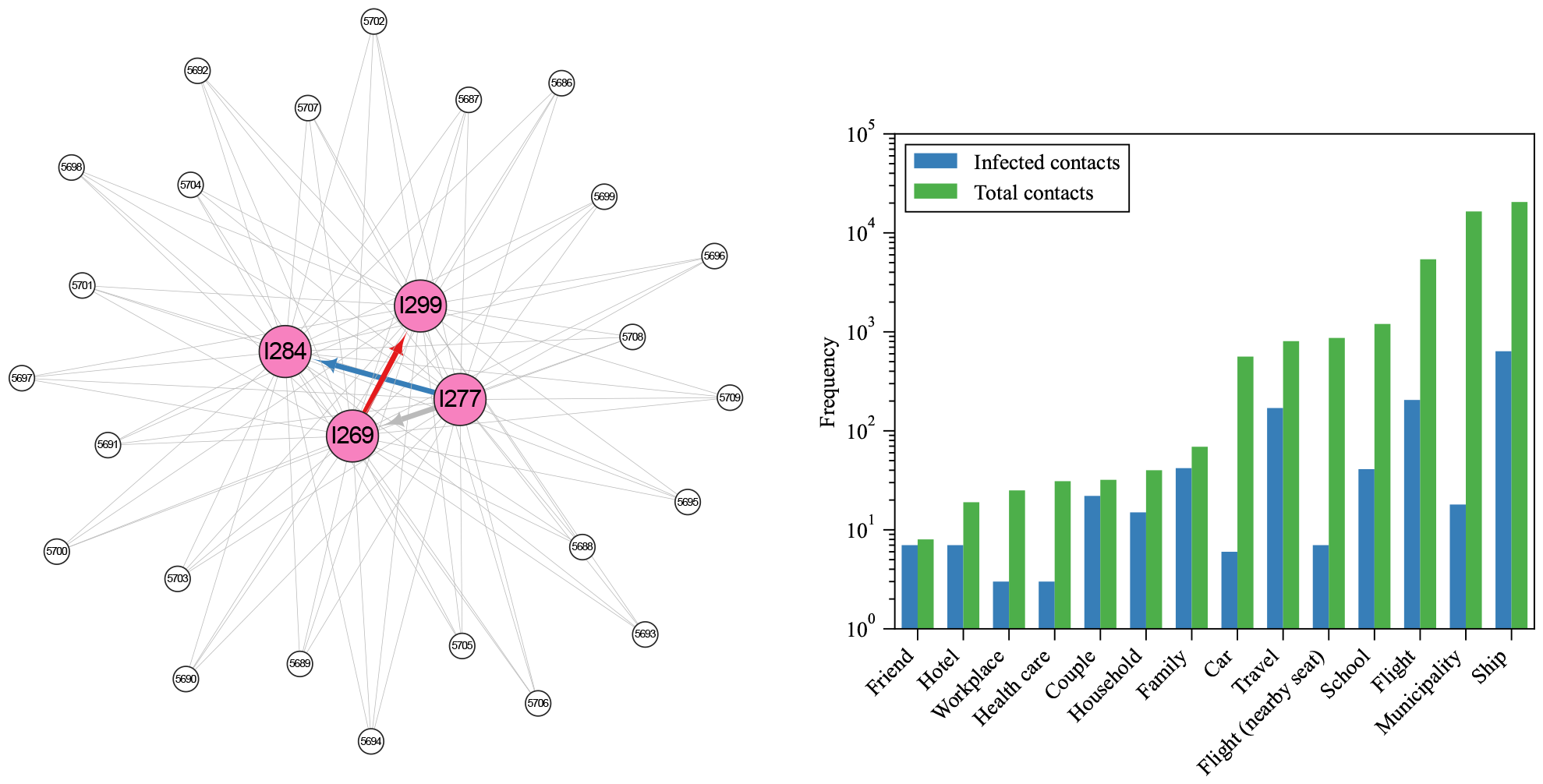
Visualization of the contact network of case I269 cluster and bar chart of each contact type. The left figure shows the isualization of the contact network of case I269 cluster. Infected cases are highlighted in pink and depicted as larger nodes, while uninfected cases are illustrated in white and represented by smaller nodes. The network is presented as a mixed graph, where infection paths are denoted by arrows and other connections are indicated by solid lines. Contact types are differentiated by color: ‘Family’ contacts are shown in red, ‘Flight’ contacts are marked in blue, and ‘Municipality’ connections are depicted in gray. The right figure shows the bar chart depicting the distribution of each contact type. The chart displays the number of edges for infected cases in blue and all edges in green. A total of 14 different contact types are represented along the x-axis.

The right panel of Figure 3 provides an overview of different contact types within the network. These contact types are categorized to ensure mutual exclusivity and comprehensive coverage. For simplicity, the plot shows larger contact groups. For example, diverse ship-related contacts, such as ‘Ship’, ‘Panshi fast combat support ship’, and ‘Coral Princess’, were consolidated under the category ‘Ship’.

## 4 Technical Validation

We conducted a thorough validation of our dataset through a crossover check using diverse datasets from various sources. Specifically, each cases’ course of disease was verified through a double-check process involving CDC press releases and press conferences, while the accuracy of the contact tracing data was confirmed by cross-referencing CDC press releases, CDC press conference, and COVID-19 Visualization. Additionally, the integrity of the final dataset underwent a double-check process involving other lab member.

To enhance the validation of our dataset, we performed an in-depth analysis and compared our results with other research focused on COVID-19 in Taiwan. Liu *et al*.^14^ conducted a study akin to ours, where they examined 321 imported cases sourced from Taiwan CDC press releases. Their analysis primarily focused on the interaction between demographic information, import sources, symptoms, and travel history. It’s essential to note that our dataset includes 579 cases, encompassing their 321 cases offering a more comprehensive view. Our data spans from January 21, 2020, to November 9, 2020, and has been meticulously transformed into a structured, accessible format, ensuring ease of access and utilization.

In our validation process, we replicated a portion of Liu *et al*.’s statistical analysis using our dataset. We isolated the 321 imported cases (case ID 1 to 373), excluding local positive cases, among which 53% were female. Notably, 37.1% fell within the 20-29 age group, and 23.7% were in the 30-39 age group, closely mirroring Liu *et al*.’s findings with only a slight difference in the 20-29 age group (37.4% in their report). Our Figure 4A reproduces Liu *et al*.’s Figure 1, depicting the age-gender distribution. Figure 4B replicates Liu *et al*.’s Figure 2, illustrating the relationship between the date of arrival and the infection source. The overall trend closely aligns with the original article, with minor variations potentially attributed to differences in defining continents or regions. When multiple travel countries were involved, and Taiwan CDC didn’t specify the infection source country, we defined the last country as the infection source. Figure 4C corresponds to their Figure 3, depicting arrival dates vs onset-of-symptom and case numbers. Our reproduction maintains the original pattern, although slight discrepancies, like those on days 1 and 2. Figure 4D replicates Liu *et al*.’s Figure 4, with minor differences due to missing information on the source of detection for some cases, especially in contact tracing.

**Figure 4.**
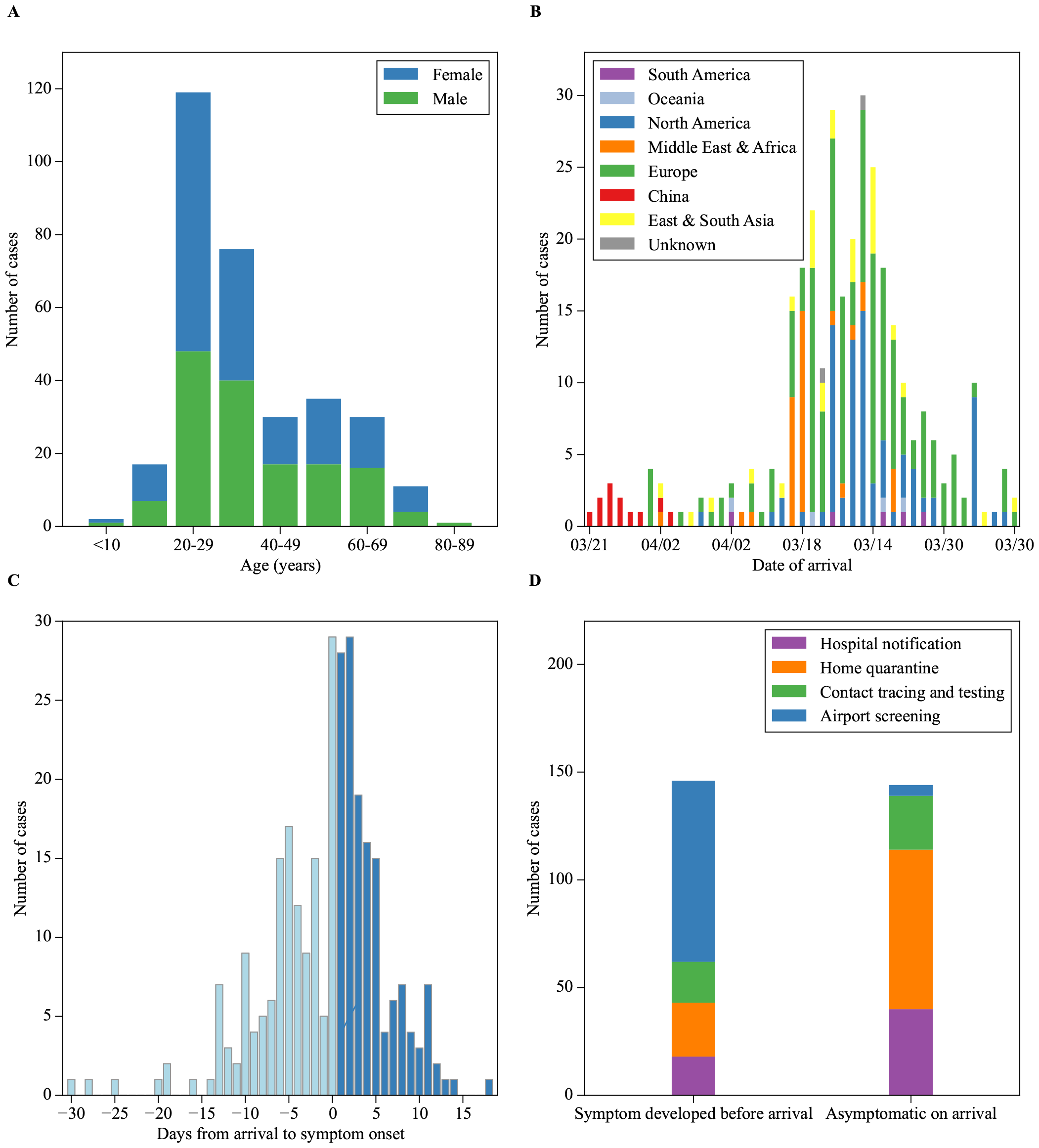
Visualization of Figures 1 to 4 from Liu *et al*. using our dataset. The overall trend aligns with their study. There are only a few minor differences. Figure B includes an additional ‘unknown’ category, indicating cases where the exact source was not identified. In Figure C, we observed more cases on day 2 than day 1, contrary to Liu *et al*.’s findings. Notably, we observed cases beyond day 13, a deviation from Liu *et al*.’s observations. In Figure D, we lack some data in the contact tracing and testing group, especially in the asymptomatic-on-arrival bar.

## 5 Usage Notes

We introduced a Taiwanese dataset that encompasses both population-level and individual-level data related to COVID-19. This meticulous approach to collecting and structuring data has resulted in a comprehensive dataset that empowers researchers to delve into individual-level COVID-19 data with confidence. To the best of our knowledge, this dataset represents the first comprehensive public structured dataset for COVID-19, providing individual-level information. However, it’s important to acknowledge the limitations of this dataset, primarily stemming from its reliance on publicly available sources. As explained by Taiwan CDC, detailed information on recovered cases has not been reported since March 12, 2020, due to privacy concerns, with only the daily count of recoveries provided. Similar limitations apply to critically ill cases. Furthermore, obtaining the precise infection date is inherently challenging and would necessitate more extensive individual contact history data. These factors collectively impact the depth of information available in our dataset. Concerning the contact information for subjects, our dataset depends on the information available in the CDC’s reports. Importantly, contact tracing poses inherent challenges, and despite the CDC’s best efforts, it is likely that some contacts may be missed. As a result, the contact numbers we provide should be considered lower-bound estimates of the true values.

In conclusion, we provided a structured dataset for COVID-19 in Taiwan in the early phase of the pandemic, covering both individual-level and population-level data. Despite the limitations due to the scope of data collection and privacy concerns, this dataset offers more details for epidemiological modelling compared to the existing population-level data. The dataset includes a wide range of variables, such as epidemiological data, course of disease data and contact types between infected and uninfected cases. We hope that this dataset will empower researchers to develop more accurate and predictive applications for combatting future pandemics. Future work could focus on expanding the dataset with more individual-level data and refining the data processing techniques to extract more valuable information.

## Data Availability

All data produced are available online at figshare "COVID-19 Taiwan data, including individual course of disease"

https://doi.org/10.6084/m9.figshare.24623964.v1

## 6 Code availability

The code for data processing and visualization can be found in the following GitHub repository: https://github.com/nordlinglab/COVID19TW-Viz

## 7 Acknowledgements

The authors acknowledgements valuable help with data confirmation from Lee Chang Jui. As non-native English speakers, we acknowledge the work of OpenAI, L.L.C. in creating GPT3.5 and GPT4, which helped us improve the readability and language of this article. We would like to thank the Ministry of Science and Technology in Taiwan for their financial support (grants number MOST 105-2218-E-006-016-MY2, 105-2911-I-006-518, 107-2634-F-006-009, 110-2222-E-006-010, and National Science and Technology Council 111-2221-E-006-186 and 112-2314-B-006-079). The funding has covered a stipend to Yu-Heng Wu and general lab expenses. The funding institution has not been involved in this study.

## 8 Author contributions statement

All authors did editing of the final manuscript. T.N. did conceptualisation, funding acquisition, methodology, project administration, resources, and supervision. Y.H.W did investigation, data curation, formal analysis, software development, and writing.

## 9 Competing interests

The authors declare no competing interests.

## Notes

### Competing Interest Statement

The authors have declared no competing interest.

### Funding Statement

This study was funded by the Ministry of Science and Technology in Taiwan (grants number MOST 105-2218-E-006-016-MY2, 105-2911-I-006-518, 107-2634-F-006-009, 110-2222-E-006-010, and National Science and Technology Council 111-2221-E-006-186 and 112-2314-B-006-079).

## References

1. Wu, Y.-H. & Nordling, T. E. M. Towards course of disease based epidemiological modelling: motivation and computational optimization. medRxiv 10.1101/2023.05.24.23290318 (2023). https://www.medrxiv.org/content/early/2023/05/28/2023.05.24.23290318.full.pdf.

2. Lee, N.-Y. et al. A case of covid-19 and pneumonia returning from macau in taiwan: Clinical course and anti-sars-cov-2 igg dynamic. J. Microbiol. Immunol. Infect. 53, 485–487 (2020).

3. Stoecklin, S. B. et al. First cases of coronavirus disease 2019 (covid-19) in france: surveillance, investigations and control measures, january 2020. Eurosurveillance 25, 2000094 (2020).

4. Garibaldi, B. T. et al. Patient trajectories among persons hospitalized for covid-19: a cohort study. Annals internal medicine 174, 33–41 (2021).

5. Wongvibulsin, S. et al. Development of severe covid-19 adaptive risk predictor (scarp), a calculator to predict severe disease or death in hospitalized patients with covid-19. Annals internal medicine 174, 777–785 (2021).

6. Lichtner, G. et al. Predicting lethal courses in critically ill covid-19 patients using a machine learning model trained on patients with non-covid-19 viral pneumonia. Sci. Reports 11, 13205 (2021).

7. Nordling, T. E. & Wu, Y.-H. Taiwan on track to end third covid-19 community outbreak. medRxiv 10.1101/2021.06.20.21259178 (2021-06-27).

8. Taiwan Centers of Disease Control. Taiwan Centers of Disease Control - News Bulletin (2021).

9. daily news, U. Visualization of contacts of taiwanese covid-19 cases (2021).

10. Regents of the National Center for High-performance Computing. COVID-19 Dashboard (2020).

11. Taiwan Centers of Disease Control. Daily Number of Cases Suspected SARS -CoV-2 Infection Tested (2021).

12. Taiwan Centers of Disease Control. Taiwan Centers for Disease Control press release (2021).

13. Wu, Y.-H. & Nordling, T. E. M. Covid-19 taiwan data, including individual course of disease, 10.6084/m9.figshare.24623964 (2024).

14. Liu, J.-Y., Chen, T.-J. & Hwang, S.-J. Analysis of imported cases of covid-19 in taiwan: a nationwide study. Int. J. Environ. Res. Public Heal. 17, 3311 (2020).

